# Scale-Up of Voluntary Medical Male Circumcisions as Part of Combination HIV Prevention Strategies – Mozambique, 2010-2023

**DOI:** 10.1101/2024.11.20.24317610

**Authors:** Marcos Canda, Lúcio Matsimbe, Nuno Gaspar, António Langa, Inácio Malimane, Jotamo Come, Daniel Chicavel

## Abstract

**Introduction:** Voluntary Medical Male Circumcision (VMMC) is a cost-effective HIV prevention strategy, reducing risk of female-to-male transmission by approximately 60%. Mozambique implemented a national VMMC program since 2010, including HIV testing to identify HIV positive men. We used program data to describe the President’s Emergency Plan for AIDS Relief (PEPFAR)-supported VMMC services in seven provinces in Mozambique.

**Methods:** We analyzed aggregate PEPFAR Monitoring, Evaluation, and Reporting data for 2010–2023 for VMMCs conducted and HIV testing at VMMC sites. VMMC coverage rate was calculated as the number of males who reported medical or traditional circumcisions over the estimated population. Test positivity rate was calculated as positive tests over the total tests conducted. Data were analyzed by fiscal year (October–September), age group (<15 years, 15–29 years, ≥30 years), and province.

**Results:** During 2010-2023 total 2,534,411 VMMCs were performed, of which, 1,032,789 (41%) among boys <15 years old, 1,368,582 (54%) among men aged 15– 29, and 133,039 (5%), men aged >30. By 2023, national VMMC coverage was 77% (higher among men aged 20–24 years, at 80%). Overall HIV test positivity rate was 1.6% (35,743 positives), with a provincial range of 0.4% to 5.9%. Older men aged 45-49 had higher HIV positivity rate of 24.3%.

**Conclusion:** VMMC implementation during twelve years reached priority boys and men aged 15-29 for immediate impact on HIV prevention. Essentially, program has circumcised a large amount of boys and men, leading to higher VMMC coverage and anticipated reductions in HIV transmission. Further, services have helped identify men already living with HIV.

## Introduction

Voluntary medical male circumcision (VMMC) is a one-time surgery that reduces the risk of female-to-male sexual transmission of HIV by 60%.^1^ In 2007, the World Health Organization and the Joint United Nations Programme on HIV and AIDS (UNAIDS) endorsed VMMC as part of combination prevention strategies to reduce new HIV infections. Starting in 2010, the U.S. President’s Emergency Plan for AIDS Relief (PEPFAR) began supporting VMMC programming in prioritized countries in southern and eastern Africa.^2^

Mozambique is an HIV high-burden country with a prevalence rate of 12.5% in 2021, corresponding to an estimated 2,460,000 people living with HIV (PLHIV) in 2023.^3^ Nationally, the HIV prevalence among adult men (aged ≥15 years) is 9.5%, corresponding to an estimated 830,000 men living with HIV.^3^

To increase VMMC as a form of HIV prevention, the Mozambique Ministry of Health (MOH), with support from PEPFAR, began staff training, program oversight, and quality assurance in 2010. PEPFAR implementing partners, along with community (for example, community lay mobilizers, faith-based organizations, public and private institutions such as sugar companies (Maragra, xinavane and Mafambisse) and institutional stakeholders (ARCUMI, AJACOMO, and the Anglican Church), support program implementation by offering the VMMC package comprised of five primary interventions: counseling about reproductive and sexual heath and risk factors for contracting HIV; HIV counseling and testing for boys and men accessing VMMC services, with referral to care and treatment services for those testing positive; VMMC procedure; post-operative care (which focus is to reduce the chance of adverse events from occurring), and health facility and community engagement.

Since 2010, the VMMC program scaled up over three phases, with geographic prioritization based on HIV incidence. In particular, provinces in the southern region (Gaza, Maputo Province and Maputo City) had higher rates of HIV and lower VMMC coverage compared to the provinces in the central and northern regions. Cabo Delgado, Inhambane, Nampula, and Niassa provinces were excluded from programming because all four provinces perform traditional circumcision and had circumcision rates of ≥80%. During phase one (October 2010–September 2014), VMMC programming prioritized southern regions and the capital cities of provinces in the central region. During phase two (October 2015– September 2019), the VMMC program rapidly expanded to most districts and health facilities from the same provinces. A greater portion of VMMC sites opened in rural areas of Zambezia, Sofala, and parts of Tete and Manica provinces. Finally, in phase three (October 2020–September 2023), VMMC programming expanded to more districts, especially in rural areas. The phased scale up of VMMC services allowed for a smooth and controlled expansion of services while assuring clients’ safety and maintenance of program quality.^2^

During the three phases of VMMC programming, the VMMC services delivery models, demand for services, and use of outreach approaches evolved. In general, VMMC programming has adapted to accommodate growing demand for VMMC services, including extended operating hours, increased staffing, multi-stakeholder collaboration, and VMMC mobile units to provide services in places with limited availability of health facilities. In phase one, services were offered at static sites and demand was spontaneous. In phase two, the key programmatic feature was the use of outreach approaches. In this phase, the VMMC programming used static, temporary, and mobile surgical units to reach clients far away from health facilities; program also used VMMC campaigns in combination with the use of mobile units in locations where there was high demand for services. Additionally, VMMC teams started conducting more active demand-creation interventions such as the use of community radio stations and community mobilizers to share information about VMMC. In phase three, the program continued to use outreach approaches for clients who had difficulty accessing VMMC. During phase one and two, all boys and men aged ≥10 years were eligible to access VMMC services. In phase three, the minimum age changed from 10 to 15 years.

This analysis describes VMMC expansion since the program’s onset in 2010, as well as HIV testing results from the VMMC program.

## Methods

Data for the VMMC program from all HF that are implementing the program were extracted from PEPFAR’s Monitoring, Evaluation, and Reporting (MER) dataset for fiscal years (FY, October–September) 2010 to 2023)^1^ Estimated circumcision coverage was sourced for 2015 IMASIDA survey and from INSIDA 2021.^3,4^

A circumcision was defined as minor surgery whereby the penis prepuce is removed by trained, accredited staff using a predefined technique. In Mozambique, two main surgical VMMC techniques are used: the dorsal slit and forceps-guide. Additionally, in 2023, the ShangRing device was piloted, toward potential scale up. Client follow-up rate was calculated as the number of clients who returned for a post-operation follow-up visit within 14 days after VMMC procedure among all clients who received VMMC services. HIV testing results were grouped in three categories: negative, positive, and indeterminate, following the national HIV testing guidelines. The HIV test positivity rate was calculated by dividing the number of clients who tested positive for HIV by total number of clients tested. We analyzed VMMC and testing data by FY, program implementation phase (one, two, three), province, and age group (<15, 15– 29, and ≥30 years).

## Results

During FY 2010–2023, 2,550,371 clients were circumcised, of which 389,221 (15%) were circumcised during phase one, 1,513,295 (60%) during phase two, and 647,855 (25%) during phase three. During this time, estimated VMMC coverage increased from 60% in 2010 to 77% in 2023. In 2023, the median provincial estimated VMMC coverage among 15-29 years old clients was 42% in Tete, Sofala was around 50%, Manica at 68%, Zambezia at 74%, Gaza at 81%, Cidade de Maputo at 88% and Maputo Province at 89%. The national estimated coverage on the same age range was 77%.

Figure 1 illustrates VMMC saturation from 2015 t0 2021 per province. VMMC saturation remained at 90% or more in northern provinces of Niassa, Cabo Delgado, and Nampula, and in the southern province of Inhambane. Circumcision in all these four provinces is performed traditionally. On the other hand, in all PEPFAR-supported provinces VMMC saturation increased in all provinces. Southern provinces of Maputo and Maputo City have reached 90% or more saturation, except Gaza province that is nearing 80%. Tete and Manica provinces are still below 50% saturation. The national VMMC saturation in 2021 was at 77% (Table 1).

**Table 1:** Number of VMMCs, by Province and Program Phases.

| Provinces | Phase 1 | Phase 2 | Phase 3 | Total VMMC | Estimated Circumcision Coverage (2021) |
| --- | --- | --- | --- | --- | --- |
| Military Mozambique | - | 122,794 | 106,697 | 229,491 | - |
| Cabo Delgado | - | 1,349 | - | 1,349 | 94% |
| Cidade De Maputo | 83,181 | 49,939 | 11,363 | 144,483 | 93% |
| Gaza | 57,819 | 152,911 | 30,183 | 240,913 | 78% |
| Manica | 24,231 | 223,878 | 124,686 | 372,795 | 47% |
| Maputo | 59,573 | 96,415 | 38,315 | 194,303 | 90% |
| Nampula | 10,922 | 3,537 | - | 14,459 | 97% |
| Sofala | 80,837 | 237,790 | 62,646 | 381,273 | 57% |
| Tete | 9,143 | 176,740 | 93,910 | 279,793 | 33% |
| Zambezia | 63,515 | 447,942 | 180,055 | 691,512 | 69% |
| **Total** | **389,221** | **1,513,295** | **647,855** | **2,550,371** | **77%** |

**Figure 1:**
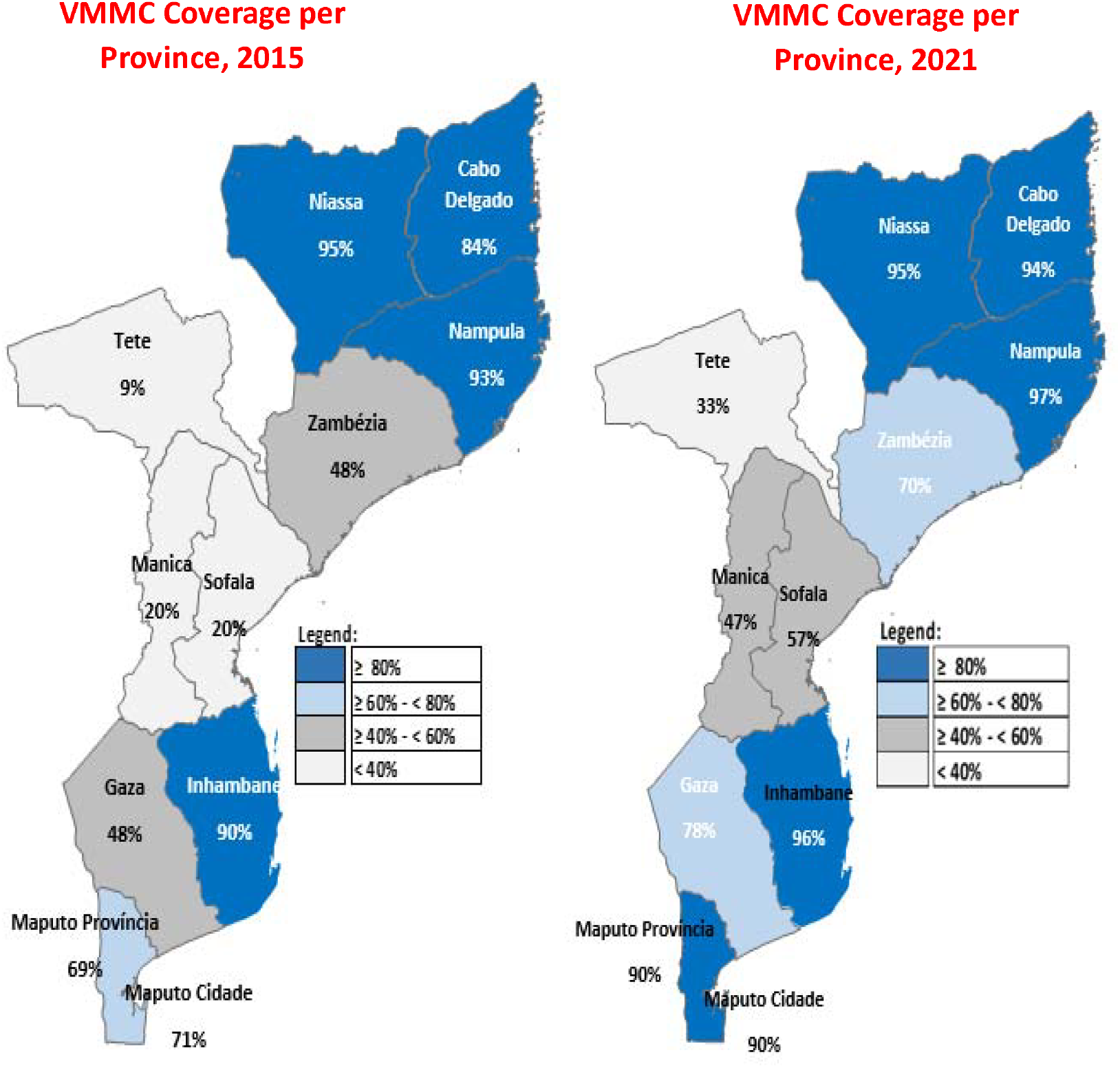
Estimated VMMC coverages per Province, 2015 and 2021 (PHIA, 2021)

Among all circumcisions performed, clients aged 15–29 years accounted for 55% (1,391,199/ 2,550,371). During phase one, clients <15 years old comprised 49% of all VMMC (192,437/389,221), 15-29 years old were 44% (171,844/389,221), and ≥30 years were 6% (24,940/389,221). In phase two, clients <15 years accounted for 47% of all VMMCs (718,025/1,513,295), 15–29 years old were 48% (720,964/1,513,295) and ≥30 years were 5% (74,306/1,513,295). In phase three, clients <15 years made up 14% of all VMMCs (89,705/647,855), 15–29 years were 77% (498,391/647,855), and ≥30 years were 9% (59,759/647,855).

Overall, the median annual circumcisions performed by province (Table 1) was 21,180 (range = 3,537 in Nampula Province to 62,865 in Zambezia Province). During phase one, the province with the lowest contribution was Tete Province (2% [9,143/389,221]) and the highest contribution was from Maputo City (21% [83,181/389,221]). During phase two, Maputo City contributed the fewest (3% [49,939/1,513,295]) and Zambezia Province contributed the most (30% [447,942/1,513,29]). During phase three, Maputo City contributed the fewest (2% [11,363/ 647,855]) and Zambezia Province contributed the most (28% [180,055/647,855]) as shown on Table 1.

During FY 2010–2023, among the 2,550,371 circumcised boys and men, 2,363,596 (93%) of clients were tested for HIV. Of these, 1,980,288 (84%) tested negative, 343,884 (14%) had indeterminate results, and 39,423 (2%) tested positive (Figure 2). HIV test positivity was lowest in Tete Province (0.4% [765/175,979]). The HIV test positivity rate has increased from 1.4% before 2020 to 2.6% in 2023. Client follow-up within 14 days after male circumcision varied between 98% and 100% across provinces and age bands in the analysis period.

**Figure 2:**
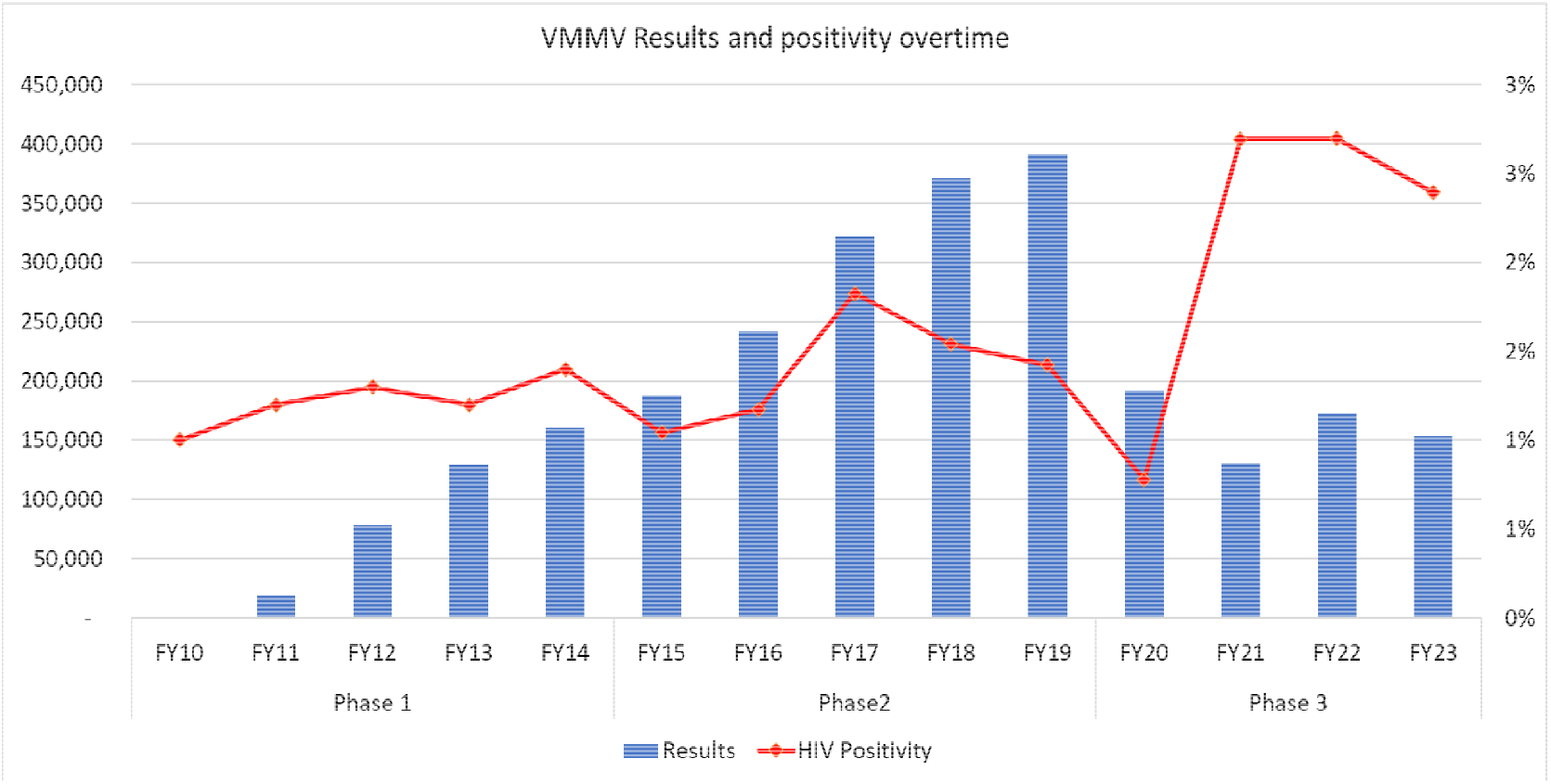
VMMC performance results and HIV positivity rates, 2010 – 2023

## Discussion

Since 2010, VMMC programming reached over 2.5 million boys and men to increase VMMC coverage to almost 80% of the eligible population in Mozambique, preventing HIV infections as a part of a combination strategy.^3^ Additionally, through HIV testing as part of the VMMC services, almost 40,000 HIV positive clients were referred to care and treatment.

Despite these impressive achievements, challenges remain. VMMC coverage varies geographically and by age group, indicating that focused efforts are needed in remote areas and among men aged 15-29 years. The disproportionate VMMC coverage across provinces and districts is likely because program scale up was performed in a phased manner.

The use of focused program implementation strategies, such as temporary VMMC campaigns to cover for the current unmet need by using geographic estimation tools can improve efficiency and facilitate reach the areas and populations with lower VMMC coverage. As well, improved data on the impact of VMMC on HIV infections averted would strengthen planning and implementation of the sustainability phase of the VMMC portfolio.

The VMMC program moved from use of a risk-screening HIV testing tool during phase 2, back to universal testing within phase 3 so that all clients should be offered an HIV test. It is believed that during the period a screening tool was used, some HIV cases could have been missed. Thus, the program returned to universal testing. We see the impact of this change in graph 2 where test positivity increases from 1.4% to 2.6%.

The VMMC program implementation involves programmatic, policy, natural and unexpected events that affect performance and accessibility of services to part of priority clients. For instance, the COVID-19 pandemic, in combination with the program minimum age shift, negatively impacted performance (graph 2) and thus, VMMC coverage at large.

There are several limitations to this analysis. We analyzed PEPFAR program data, which are subject to reporting and data entry errors, despite continual data quality assurance exercises. Program data also cannot be used to directly assess VMMC coverage or estimate HIV infections averted. Finally, these data do not capture circumcisions performed outside of PEFPAR-supported VMMC sites.

## Conclusion

Through this review, and despite some challenges, VMMC services in Mozambique continue to contribute to HIV prevention in combination with other prevention strategies, supports case identification efforts and continue to focus on areas where VMMC coverage remains low.

## Data Availability

All data produced are available from PEPFAR Monitoring, Evaluation, and Reporting program data.

https://data.pepfar.gov/datasets#PDD

## Conflict of Interest

all authors declare no competing interests.

## Funding Acknowledgement

This manuscript has been supported by the President’s Emergency Plan for AIDS Relief (PEPFAR), through the Centers for Disease Control and Prevention (CDC), The findings and conclusions in this manuscript are those of the author(s) and do not necessarily represent the official position of the funding agencies (CDC, OGAC/PEPFAR).

## Footnotes

1 This activity was reviewed by CDC, deemed not research, and was conducted consistent with applicable federal law and CDC policy. See e.g., 45 C.F.R. part 46.102(I)(2), 21 C.F.R. part 56; 42 U.S.C. 241(d); 5 U.S.C. 552a; 44 U.S.C. 3501 et seq.

## Notes

### Competing Interest Statement

The authors have declared no competing interest.

### Funding Statement

This manuscript has been supported by the President's Emergency Plan for AIDS Relief (PEPFAR) through the U.S. Centers for Disease Control and Prevention (CDC). The findings and conclusions in this manuscript are those of the author(s) and do not necessarily represent the official position of the funding agencies.

### Author Declarations

The study used PEPFAR Monitoring, Evaluation, and Reporting program.

### Summary of Updates

VMMC coverage maps have been revised and updated to align with correct legend colors and remove decimal units.

